# Referral pathways, ETAT triage acuity, and inpatient outcomes among children presenting to a national tertiary paediatric emergency unit in Ghana: a prospective cohort study

**DOI:** 10.64898/2026.06.17.26355891

**Authors:** William Kojo Ansah Obeng, Lorna Awo Renner, Yakubu Alhassan, Tom Akuetteh Ndanu, Bamenla Quarm Goka

**Affiliations:** Department of Child Health, Korle Bu Teaching Hospital; Department of Child Health, University of Ghana Medical School, College of Health Sciences, University of Ghana; Department of Biostatistics, School of Public Health, University of Ghana; Department of Community and Preventive Dentistry, University of Ghana Dental School, College of Health Sciences, University of Ghana

**Author notes:** Corresponding author: Dr William KA Obeng;, Department of Child Health, Korle Bu Teaching Hospital, P.O. Box KB 77, Korle Bu, Accra, Ghana.

**Keywords:** referral systems, ETAT, triage, paediatric emergency, Ghana, LMIC, mortality, cohort study

## Abstract

Emergency referral systems in sub-Saharan Africa are fragmented, and children reaching tertiary facilities through different referral pathways often arrive in advanced clinical states. Prospective data simultaneously characterising referral patterns, triage acuity at presentation, diagnostic case mix, and inpatient mortality at a national tertiary paediatric emergency unit are lacking from West Africa.

This prospective cohort study enrolled 675 consecutively presenting children aged one month to 12 years at the Paediatric Emergency Unit of Korle Bu Teaching Hospital, Accra, Ghana, from February to December 2019. The primary outcome was all-cause inpatient mortality. Key variables collected included referral status and facility tier, Emergency Triage Assessment and Treatment (ETAT) triage category, ICD-10 diagnostic classification, Oyedeji socioeconomic classification, and time from symptom onset to PEU registration. Crude odds ratios were computed for all candidate predictors. Multivariable logistic regression was conducted using complete case analysis (*n* = 613).

Of 675 children, 63.0% (*n* = 425) were referred from another health facility; referred children had higher ETAT emergency triage category rates than self-presenting children (32.7% vs 27.6%, *p* < 0.001). Overall inpatient mortality was 9.9% (67/675). Mortality varied by referral source: 16.7% among secondary/regional hospital referrals, 11.0% among lower-tier facility referrals (district, municipal, CHAG, polyclinic, private, health centre, and maternity home facilities combined, *n* = 356), 7.6% among self-presenting children, and 7.4% among tertiary referrals. Overall, 30.8% of children were classified as ETAT emergencies on arrival, with case fatility rate of 21.6%. The three most common diagnostic domains were respiratory conditions (17.2%), blood and haematological disorders (17.0%), and digestive presentations (16.4%). Inpatient mortality was highest in neoplastic disease (33.3%, *n* = 30) and circulatory presentations (31.0%, *n* = 29). In the primary multivariable analysis (*n* = 613, 51 events; events-per-variable ratio 4.2), no referral tier was independently associated with inpatient mortality after adjustment. Referral from secondary/regional hospitals showed a borderline non-significant association (adjusted odds ratio 3.09, 95% CI 0.96 to 9.90, *p* = 0.058). School going children (60–119 months) had higher odds of inpatient death than infants (adjusted odds ratio 5.56, 95% CI 1.16 to 26.53, *p* = 0.032), as did adolescents (adjusted odds ratio 10.01, 95% CI 2.15 to 46.69, *p* = 0.003). ETAT emergency category and lower socioeconomic status were not independently significant in this model. A pre-specified sensitivity analysis using the full analytic cohort (*n* = 674, events-per-variable ratio 6.7) with collapsed referral categories did not confirm any referral tier association; ETAT emergency category and lower SES were independently associated in the sensitivity model. All multivariable estimates should be regarded as exploratory.

This prospective cohort provides simultaneous characterisation of referral patterns, ETAT triage acuity, diagnostic case mix, and inpatient mortality at a national tertiary paediatric emergency unit in West Africa. The referral-mortality gradient and high ETAT emergency category proportion document the severity of illness arriving through different referral pathways at this facility. The association between secondary/regional hospital referral and inpatient mortality is hypothesis-generating and requires replication in an adequately powered multicentre study before any service-level conclusions can be drawn.

## INTRODUCTION

Child mortality in sub-Saharan Africa (SSA) remains high. In Ghana, the under-five mortality rate was approximately 44 per 1,000 live births in 2021, based on UNICEF estimates. [1] Emergency care system gaps have been documented as contributors to preventable infant and child deaths in this setting. [2,3] A study of hospitalised children in Ghana documented a co-existing burden of communicable and non-communicable disease presentations, consistent with an ongoing epidemiological transition in paediatric inpatient populations. [4] At tertiary facilities in SSA, a substantial proportion of paediatric inpatient deaths occur within the first 24 hours of admission. [5,6] This pattern suggests that many children arrive in advanced clinical states, and that the quality of triage, stabilisation, and referral before reaching tertiary care may contribute to outcomes at the receiving facility. [7]

Referral systems in SSA are characterised by fragmented pathways, limited pre-referral stabilisation capacity at lower-tier facilities, and bypassing of intermediate levels of care. [8,9,24] In Ghana, children presenting to a national tertiary paediatric emergency unit may arrive without a referral or following referral from a primary health centre, district or municipal hospital, secondary/regional hospital, polyclinic, maternity home, private facility, or Christian Health Association of Ghana (CHAG) institution. [10] Arrival acuity and the risk of inpatient death may differ by referral source, but prospective data on this relationship are scarce in West Africa. Referral quality research in the region has focused primarily on surgical and elective care populations rather than paediatric emergency presentations. [11]

The Emergency Triage Assessment and Treatment (ETAT) framework, which classifies children as emergency, priority, or queue on arrival, is widely implemented in paediatric emergency units across SSA. [12] It was designed to identify children requiring immediate resuscitation and to standardise the initial clinical response. Training coverage at primary care level remains low; one study from Malawi documented ETAT training in fewer than 12% of primary care workers. [13] Children referred from lower-tier facilities may therefore arrive at tertiary units without prior ETAT assessment or pre-referral stabilisation. [13] ETAT acuity recorded at the receiving centre may in this context reflect not only the child’s underlying disease severity but also the quality of the referral process.

Prospective cohort data from West Africa that simultaneously describe referral patterns, ETAT triage acuity, diagnostic case mix, and inpatient mortality at a national tertiary paediatric emergency unit are lacking. This study uses a prospective cohort of 675 children presenting to the Paediatric Emergency Unit (PEU) of Korle Bu Teaching Hospital (KBTH), Ghana’s national tertiary referral centre, to address that gap. The study aimed to describe referral patterns and triage acuity across facility tiers, to characterise the diagnostic case mix and its associated inpatient mortality, and to identify factors associated with inpatient mortality in an exploratory multivariable analysis.

## METHODS

### Study design and setting

We conducted a prospective cohort study at the Paediatric Emergency Unit (PEU) of Korle Bu Teaching Hospital (KBTH), Accra, Ghana, from February to December 2019. KBTH is the principal national tertiary referral hospital in Ghana, serving all 16 administrative regions. The PEU operates as a general paediatric emergency unit and is the emergency intake for the full range of paediatric subspecialty services within the Department of Child Health (DCH).

### Study population

Children aged one month to 12 years presenting consecutively to the PEU during the study period were eligible. Exclusion criteria were: refusal of consent; incomplete outcome data; caregiver abscondment before assessment; and death within one hour of arrival. In total, 675 children were enrolled; all 675 had complete ETAT triage data and are included in all descriptive and unadjusted analyses.

### Data collection

Structured questionnaires were administered at PEU registration by trained research assistants who had no role in clinical management. Variables collected included: age (months), sex, socioeconomic status (Oyedeji 1985 occupational and educational classification, class I–II upper, class III middle, class IV–V lower), [14] referral status (referred from another facility vs self-presented), referring facility level, time from symptom onset to PEU registration (hours), ETAT triage category at arrival (emergency, priority, or queue), ICD-10 primary diagnostic classification, and inpatient outcomes.

Referring facility levels reflect Ghana’s tiered public health system. Tertiary hospitals provide national specialist care. Secondary/regional hospitals serve administrative regions. District and municipal hospitals provide first-level inpatient care across district and municipal catchment areas. Polyclinics are government-operated sub-district outpatient facilities, and health centres are smaller community-level primary care facilities. Christian Health Association of Ghana (CHAG) facilities are a faith-based provider network operating at all levels of the health system, from community clinics to hospitals. Private hospitals are independent not-for-profit and for-profit facilities. Maternity homes provide primarily obstetric services.

The Oyedeji classification was developed in Nigeria and has not been validated in Ghana. No validated Ghanaian socioeconomic classification for paediatric clinical research has been identified in the published literature; its use here follows convention in West African paediatric hospital-based studies.

### Outcome measures

The primary outcome was all-cause inpatient mortality, defined as death occurring at any point during the hospital episode. Secondary outcomes were discharge destination and length of stay.

### Statistical analysis

Continuous variables are described as median and interquartile range; categorical variables as frequencies and percentages. Chi-square tests were used for group comparisons of categorical variables. Crude odds ratios (cOR) with 95% confidence intervals (CI) were calculated for all candidate predictors and are presented for the full cohort (*n* = 675) in Table 1 for descriptive purposes. Variables with *p* < 0.2 in bivariable analysis within the complete case dataset and variables of established clinical relevance to the study objectives were entered into a multivariable logistic regression model to identify factors independently associated with inpatient mortality.

**Table 1.** Participant characteristics, referral source distribution, and case fatality rates (*n* = 675). CFR, case fatality rate; cOR, crude odds ratio; CI, confidence interval; CHAG, Christian Health Association of Ghana; ETAT, Emergency Triage Assessment and Treatment. cOR calculated relative to the reference category shown within each variable section. †Health centre and maternity home referrals had zero mortality events; OR not calculable. cOR values were computed in IBM SPSS Statistics version 23. Age groups reflect the Age_category1 variable in the submitted dataset; the manuscript text references age groups derived from continuous age in months (see Data Availability Statement). ‡ Sex data were missing for 6 participants, 1 of whom died. Deaths in the male and female rows therefore sum to 66, not 67.

| Variable and category | *n* | Deaths | CFR (%) | cOR (95% CI) full cohort *n* = 675 | *p* |
| --- | --- | --- | --- | --- | --- |
| **Referral source** |  |  |  |  |  |
| **Self-presented** | 250 | 19 | 7.6 | 1.00 (ref) | — |
| Secondary/regional hospital | 42 | 7 | 16.7 | 2.43 (0.95–6.20) | 0.055 |
| District/municipal hospital | 125 | 21 | 16.8 | 2.45 (1.27–4.76) | 0.008 |
| CHAG facility | 42 | 5 | 11.9 | 1.64 (0.58–4.67) | 0.344 |
| Polyclinic | 78 | 7 | 9.0 | 1.20 (0.48–2.97) | 0.694 |
| Health centre | 7 | 0 | 0.0 | N/A† | — |
| Maternity home | 3 | 0 | 0.0 | N/A† | — |
| Private hospital | 101 | 6 | 5.9 | 0.77 (0.30–1.98) | 0.587 |
| Tertiary institution | 27 | 2 | 7.4 | 0.97 (0.21–4.42) | 0.971 |
| **ETAT triage category** |  |  |  |  |  |
| **Queue/non-urgent** | 51 | 3 | 5.9 | 1.00 (ref) | — |
| Priority | 416 | 19 | 4.6 | 0.77 (0.22–2.68) | 0.675 |
| Emergency | 208 | 45 | 21.6 | 4.42 (1.31–14.85) | 0.024 |
| **Age group** |  |  |  |  |  |
| **Infant (reference)** | 140 | 14 | 10.0 | 1.00 (ref) | — |
| Preschool (1–5 years) | 243 | 20 | 8.2 | 0.81 (0.39–1.65) | 0.557 |
| School going (5–9 years) | 177 | 15 | 8.5 | 0.83 (0.39–1.79) | 0.640 |
| Adolescent (10–12 years) | 115 | 18 | 15.7 | 1.67 (0.79–3.52) | 0.179 |
| **Socioeconomic status (Oyediji)** |  |  |  |  |  |
| **Upper class (classes I–II)** | 118 | 4 | 3.4 | 1.00 (ref) | — |
| Middle class (class III) | 81 | 3 | 3.7 | 1.10 (0.24–5.03) | 0.906 |
| Lower class (classes IV–V) | 475 | 60 | 12.6 | 4.12 (1.47–11.58) | 0.024 |
| **Sex‡** |  |  |  |  |  |
| **Male** | 371 | 38 | 10.2 | 1.00 (ref) | — |
| Female | 298 | 28 | 9.4 | 0.91 (0.54–1.54) | 0.726 |

One participant had missing continuous age in months; age category was determinable from caregiver report and this participant is retained in all analyses. Data were missing for sex (*n* = 6) and socioeconomic status (*n* = 1). The participant with missing socioeconomic status was excluded from the complete case analysis; those with missing sex data were retained in all other analyses.

Collinearity was assessed using variance inflation factors (VIF) computed from the design matrix. All VIF values were below the conventional threshold of 5. Full values for all 12 dummy terms are provided in S2. The highest values were for the ETAT category dummy terms (maximum 3.97). These reflect the structural negative correlation between dummy variables encoding the same three-level categorical variable, not collinearity with other predictors in the model. The structural minimum for ETAT dummies derived from the ETAT marginal distribution alone was 3.61. The observed value of 3.97 is consistent with small additional cross-predictor correlations. VIF values for all other dummy terms ranged from 1.07 to 2.16. No problematic collinearity was identified.

Clinically plausible two-way interaction terms (ETAT category with referral tier, age group, and socioeconomic status; age group with socioeconomic status; and referral tier with socioeconomic status) were evaluated using likelihood ratio tests; none reached statistical significance (all *p* > 0.20) and no interaction terms were retained in the final model.

Predictors in the final model were ETAT category (three levels; reference: queue), referral facility level (six groups; reference: self-presented), age group (four levels; reference: infant ≤12 months), and socioeconomic status (three levels; reference: upper). Referral facility types were partially combined according to Ghana’s health system tier structure. Polyclinics, health centres, and maternity homes are sub-district primary care facilities serving as the initial point of formal health contact before district-level hospitals; [15] they were grouped as one primary care tier. District and municipal hospitals were grouped with CHAG facilities because CHAG hospitals predominantly operate at the district hospital level, providing first-level inpatient care. [16] Entering all eight individual facility types as separate predictors would have required 14 model parameters (EPV=3.6); partial grouping based on these clinical tier distinctions reduced this to 12 parameters (EPV=4.2). Results are reported as adjusted odds ratios (aOR) with 95% CI. Statistical significance was set at *p* < 0.05. Time from symptom onset to PEU registration was recorded only for referred children and was excluded from the multivariable model; multiple imputation was not performed. Analyses were performed in IBM SPSS Statistics version 23 (IBM Corp., Armonk, NY, USA) and Stata IC version 16 (StataCorp., College Station, TX, USA). This study is reported in accordance with the STROBE guidelines for cohort studies; the completed checklist is submitted as a supplementary file.

### Ethical approval

Ethical approval was granted by the KBTH Scientific and Technical Committee and Institutional Review Board (KBTH-IRB 00073/2017, with extension). Written informed consent was obtained from caregivers. Child assent was obtained from children aged seven years and above who were oriented at the time of assessment. The study was conducted in accordance with the Declaration of Helsinki.

### Companion manuscript declaration

This manuscript examines referral patterns, ETAT triage acuity, diagnostic case mix, and inpatient mortality in the full enrolled cohort (*n* = 675). Three companion manuscripts from the same prospective cohort, reporting PEWS-based analyses, are currently under review. Each addresses a distinct, non-overlapping research question. Full details of the companion manuscripts and a formal non-duplication statement are provided in the Declarations section.

## RESULTS

### Participant characteristics

In total, 675 children were enrolled. The median age was 48 months (IQR 16–96 months). Of these, 55.0% (*n* = 371) were male. By age group: infants (≤12 months) 20.7% (*n* = 140), preschool children (1–5 years) 36.0% (*n* = 243), school-going children (5–9 years) 26.2% (*n* = 177), and adolescents (10–12 years) 17.0% (*n* = 115). Most children were from lower socioeconomic groups (Oyedeji classes IV–V, 70.4%, *n* = 475); 12.0% (*n* = 81) were middle class and 17.5% (*n* = 118) upper class. Full participant characteristics are shown in Table 1.

### Referral source distribution and case fatality rates

Of 675 children, 63.0% (*n* = 425) were referred from another health facility; 37.0% (*n* = 250) self-presented without prior referral. Among referred children, the most common referral source was district and municipal hospitals (29.4%, *n* = 125), followed by private hospitals (23.8%, *n* = 101), polyclinics (18.4%, *n* = 78), secondary/regional hospitals (9.9%, *n* = 42), CHAG facilities (9.9%, *n* = 42), tertiary institutions (6.4%, *n* = 27), health centres (1.6%, *n* = 7), and maternity homes (0.7%, *n* = 3). Referral pathway, ETAT triage distribution, and inpatient outcome data are summarised in Fig 1; individual referral source data and mortality are shown in Table 1.

**Fig 1.**
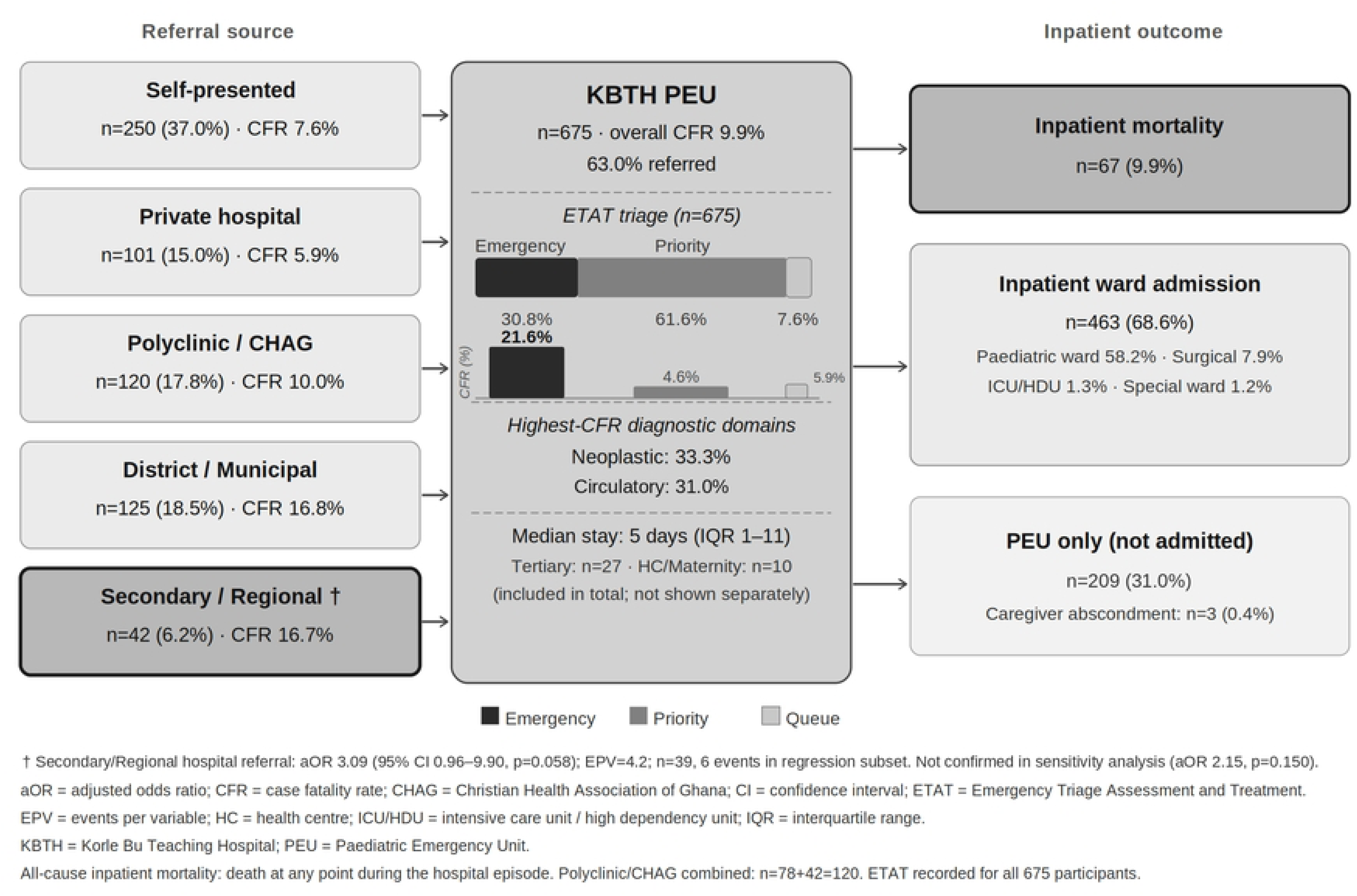
Referral pathways, ETAT triage acuity, and inpatient outcomes at KBTH PEU, Ghana, 2019. Flow diagram showing the referral source distribution, ETAT triage acuity on arrival, and inpatient outcome categories among 675 children presenting to the Paediatric Emergency Unit (PEU), Korle Bu Teaching Hospital, Accra, Ghana, February–December 2019. Referral sources are grouped by facility tier with case fatality rates (CFR). ETAT triage acuity is shown as a proportional stacked bar (dark grey = Emergency; mid grey = Priority; light grey = Queue). The Secondary/Regional hospital box (darker shading, bold border) had the highest unadjusted and adjusted mortality among referred groups. Health centre and maternity home referrals (*n* = 10 combined) and tertiary referrals (*n* = 27) are included in the PEU total but not shown as separate source boxes. † Secondary/Regional hospital: aOR 3.09 (95% CI 0.96–9.90), p = 0.058 in primary analysis (*n* = 613, EPV = 4.2); borderline non-significant. Not confirmed in sensitivity analysis (aOR 2.15, *p* = 0.150, *n* = 674, EPV = 6.7). All estimates are exploratory. CFR = case fatality rate; CHAG = Christian Health Association of Ghana; ETAT = Emergency Triage Assessment and Treatment; IQR = interquartile range; PEU = Paediatric Emergency Unit.

Unadjusted case fatality rates (CFR) varied by referral source. District and municipal hospital referrals had the highest rate at 16.8% (21/125), closely followed by secondary/regional hospital referrals at 16.7% (7/42), CHAG facility referrals at 11.9% (5/42), polyclinic referrals at 9.0% (7/78), self-presenting children at 7.6% (19/250), tertiary referrals at 7.4% (2/27), and private hospital referrals at 5.9% (6/101). Health centre and maternity home referrals recorded no deaths (0/7 and 0/3 respectively). Fractions show deaths/total in each group. Event counts were small in several categories; individual CFR should be interpreted cautiously. These figures are unadjusted; the multivariable analysis below accounts for potential confounders. When referral sources were grouped by tier, the CFR was 16.7% (7/42) among secondary/regional hospital referrals, 11.0% (39/356) among lower-tier referrals (district, municipal, CHAG, polyclinic, private, health centre, and maternity home facilities combined), 7.6% (19/250) among self-presenting children, and 7.4% (2/27) among tertiary referrals.

### ETAT triage acuity and case fatality rates

ETAT triage category was recorded for all 675 participants. Of these, 30.8% (*n* = 208) were ETAT emergency, 61.6% (*n* = 416) priority, and 7.6% (*n* = 51) queue/non-urgent. Among referred children (*n* = 425), 32.7% were ETAT emergency compared with 27.6% among self-presenting children (*n* = 250; chi-square *p* < 0.001 for overall ETAT distribution by referral status).

Inpatient mortality differed by ETAT category: 21.6% (45/208) in the emergency category, 4.6% (19/416) in the priority category, and 5.9% (3/51) in the queue category. The higher CFR in the queue group compared with priority reflects small event counts (3 deaths) and should be interpreted with caution. Emergency classification carried a cOR of 4.42 (95% CI 1.31 to 14.85, *p* = 0.024) relative to queue. Priority was not significantly different from queue (cOR 0.77, 95% CI 0.22 to 2.68, *p* = 0.675).

### Diagnostic case mix and inpatient mortality

The three most common diagnostic domains were respiratory conditions (17.2%, *n* = 116), blood and blood-forming organ disorders (17.0%, *n* = 115), and digestive conditions (16.4%, *n* = 111). Inpatient mortality was highest in neoplastic disease (33.3%, 10/30) and circulatory presentations (31.0%, 9/29). ETAT emergency classification was most frequent in respiratory conditions, congenital abnormalities, and nervous system presentations; domain-level ETAT data are shown in Table 2.

**Table 2.** ICD-10 diagnostic domain distribution, ETAT triage acuity, and case fatality rates (*n* = 675). CFR, case fatality rate; ETAT, Emergency Triage Assessment and Treatment; ICD-10, International Classification of Diseases, 10th revision. Row percentages for ETAT columns show the proportion of each domain classified in that triage category. Domains with zero CFR had no deaths during the study period.

| ICD-10 diagnostic domain | *n* | ETAT<br>Emergency<br>*n* (%) | ETAT<br>Priority<br>*n* (%) | ETAT<br>Queue<br>*n* (%) | Deaths<br>*n* | CFR<br>(%) |
| --- | --- | --- | --- | --- | --- | --- |
| Respiratory | 116 | 71 (61%) | 39<br>(34%) | 6 (5%) | 11 | 9.5% |
| Blood and blood-forming organs | 115 | 16 (14%) | 99<br>(86%) | 0 (0%) | 4 | 3.5% |
| Digestive | 111 | 13 (12%) | 84<br>(76%) | 14<br>(13%) | 6 | 5.4% |
| Genitourinary | 67 | 12 (18%) | 46<br>(69%) | 9<br>(13%) | 8 | 11.9% |
| Nervous system | 58 | 28 (48%) | 24<br>(41%) | 6<br>(10%) | 7 | 12.1% |
| Certain infectious diseases | 46 | 17 (37%) | 27<br>(59%) | 2 (4%) | 5 | 10.9% |
| Neoplastic disease | 30 | 2 (7%) | 24<br>(80%) | 4<br>(13%) | 10 | 33.3% |
| Circulatory | 29 | 16 (55%) | 13<br>(45%) | 0 (0%) | 9 | 31.0% |
| Congenital abnormalities | 26 | 15 (58%) | 11<br>(42%) | 0 (0%) | 3 | 11.5% |
| Symptoms and signs | 25 | 6 (24%) | 17<br>(68%) | 2 (8%) | 3 | 12.0% |
| Ear and mastoid | 16 | 7 (44%) | 5 (31%) | 4<br>(25%) | 0 | 0.0% |
| Endocrine/nutritional/metabolic | 10 | 2 (20%) | 8 (80%) | 0 (0%) | 1 | 10.0% |
| Injury and poisoning | 9 | 2 (22%) | 7 (78%) | 0 (0%) | 0 | 0.0% |
| Skin and subcutaneous tissue | 9 | 0 (0%) | 7 (78%) | 2<br>(22%) | 0 | 0.0% |
| Musculoskeletal | 6 | 1 (17%) | 3 (50%) | 2<br>(33%) | 0 | 0.0% |
| Mental and behavioural | 2 | 0 (0%) | 2<br>(100%) | 0 (0%) | 0 | 0.0% |
| **Total** | **675** | **208**<br>**(30.8%)** | **416**<br>**(61.6%)** | **51**<br>**(7.6%)** | **67** | **9.9%** |

### Presentation timing by triage category and inpatient mortality among referred children

Time from symptom onset to PEU registration was recorded for all 425 referred children (63.0% of the total cohort). For referred children, this interval encompasses time at the referring facility before transfer and is not equivalent to pre-healthcare-seeking delay. The median was 2 hours (IQR 1–6 hours). Of referred children, 82.1% (*n* = 349) arrived within 24 hours of symptom onset, 14.6% (*n* = 62) between 24 and 72 hours, and 3.3% (*n* = 14) beyond 72 hours.

ETAT emergency classification was highest among referred children presenting within 24 hours of symptom onset (34.7%, 121/349), compared with 25.8% (16/62) among those presenting between 24 and 72 hours, and 14.3% (2/14) beyond 72 hours; this difference was not statistically significant (chi-square *p* = 0.292). Inpatient mortality did not differ significantly by time category: 10.9% (38/349) for presentations within 24 hours, 14.5% (9/62) for 24 to 72 hours, and 7.1% (1/14) beyond 72 hours (chi-square *p* = 0.625). The > 72h group was small (*n* = 14); estimates for this group should be interpreted with caution.

### Inpatient outcomes

Overall inpatient mortality was 9.9% (67/675). In total, 68.6% (*n* = 463) were admitted to an inpatient ward. Discharge destinations were: paediatric medical ward 58.2% (*n* = 393), paediatric surgical ward 7.9% (*n* = 53), ICU/HDU 1.3% (*n* = 9), and special ward 1.2% (*n* = 8). 31.0% (*n* = 209) remained in the PEU without ward admission, and caregiver abscondment was recorded in 3 cases (0.4%). Median length of stay was 5 days (IQR 1–11).

### Multivariable analysis

The multivariable analysis included 613 participants with complete data for all model predictors (51 mortality events; Table 3). The six referral categories reflect Ghana’s health system tier structure; entering all eight individual facility types separately would have required 14 model parameters (EPV=3.6) rather than the 12 used (EPV=4.2). No referral tier was independently associated with inpatient mortality after adjustment. Referral from secondary/regional hospitals showed a borderline non-significant association compared with self-presentation (aOR 3.09, 95% CI 0.96 to 9.90, *p* = 0.058), based on a small subgroup (*n* = 39, 6 mortality events). Referral from district/municipal/CHAG facilities was not independently significant (aOR 2.03, 95% CI 0.89 to 4.61, *p* = 0.091). Referral from polyclinic, health centre, and maternity home (aOR 0.95, 95% CI 0.31 to 2.95, *p* = 0.930), private hospital (aOR 0.84, 95% CI 0.27 to 2.64, *p* = 0.766), and tertiary facilities (aOR 2.64, 95% CI 0.51 to 13.68, *p* = 0.248) were not independently associated with mortality.

**Table 3.** Multivariable logistic regression: primary model (*n* = 613) and sensitivity model (*n* = 674). aOR, adjusted odds ratio; CI, confidence interval; CHAG, Christian Health Association of Ghana; ETAT, Emergency Triage Assessment and Treatment; EPV, events per variable; HC, health centre. Primary model: *n* = 613, 51 mortality events, EPV = 4.2; complete-case analysis. Sensitivity model: *n* = 674, 67 mortality events, EPV = 6.7; collapsed referral categories. AUROC: primary model 0.804 (95% CI 0.742–0.859), sensitivity model 0.786 (95% CI 0.734–0.836). Lower tier (combined) appears only in the sensitivity model. All estimates are exploratory; EPV below 10 in both models limits precision of adjusted estimates. Participant counts in this table reflect the primary complete-case subset (*n* = 613) and differ from Table 1 (*n* = 675); the difference of 62 is explained in the Strengths and Limitations section.

| Variable and category | *n*<br>(primary<br>*n* = 613) | Deaths<br>(primary) | aOR (95% CI)<br>primary model | *p* | aOR (95% CI)<br>sensitivity *n* =<br>674 |
| --- | --- | --- | --- | --- | --- |
| **Referral facility level** |  |  |  |  |  |
| *Self-presented* (reference) | — | — | 1.00 | — | 1.00 |
| Secondary/regional hospital | 39 | 6 | 3.09 (0.96–9.90) | 0.058 | 2.15 (0.76–6.07)<br>*p* = 0.150 |
| District/municipal/CHAG | 155 | 20 | 2.03 (0.89–4.61) | 0.091 | Included in<br>lower tier |
| Polyclinic/HC/maternity home | 84 | 5 | 0.95 (0.31–2.95) | 0.930 | Included in<br>lower tier |
| Private hospital | 91 | 5 | 0.84 (0.27–2.64) | 0.766 | Included in<br>lower tier |
| Tertiary institution | 26 | 2 | 2.64 (0.51–13.68) | 0.248 | 1.99 (0.40–9.83)<br>*p* = 0.399 |
| Lower tier (combined) | — | — | — | — | 1.19 (0.63–2.25)<br>*p* = 0.590 |
| **Age group** |  |  |  |  |  |
| *Infant ≤12 months* (reference) | 94 | 2 | 1.00 | — | 1.00 |
| Preschool 13–60 months | 235 | 19 | 4.21 (0.92–19.17) | 0.063 | 0.72 (0.34–1.56)<br>*p* = 0.409 |
| School going 60–119 months | 171 | 13 | 5.56 (1.16–26.53) | 0.032 | 1.03 (0.45–2.33)<br>*p* = 0.948 |
| Adolescent ≥120 months | 113 | 17 | 10.01 (2.15–46.69) | 0.003 | 1.92 (0.86–4.29)<br>*p* = 0.113 |
| **ETAT triage category** |  |  |  |  |  |
| *Queue/non-urgent* (reference) | 43 | 2 | 1.00 | — | 1.00 |
| Priority | 396 | 16 | 0.52 (0.10–2.60) | 0.426 | 0.57 (0.15–2.17)<br>*p* = 0.410 |
| Emergency | 174 | 33 | 3.60 (0.75–17.35) | 0.110 | 3.82 (1.04–13.97) *p* = 0.043 |
| **Socioeconomic status (Oyedeji)** |  |  |  |  |  |
| *Upper class (classes I–II)*<br>(reference) | 104 | 3 | 1.00 | — | 1.00 |
| Middle class (class III) | 74 | 2 | 0.83 (0.13–5.44) | 0.845 | 1.23 (0.26–5.89)<br>*p* = 0.795 |
| Lower class (classes IV–V) | 435 | 46 | 3.03 (0.88–10.50) | 0.080 | 4.30 (1.49–12.44) *p* = 0.007 |

Older age was independently associated with inpatient mortality. Children in the school going age group (60–119 months) had higher odds of death than infants (aOR 5.56, 95% CI 1.16 to 26.53, *p* = 0.032), and adolescents (≥ 120 months) had the highest odds (aOR 10.01, 95% CI 2.15 to 46.69, *p* = 0.003). Confidence intervals were wide, reflecting the small infant reference group (*n* = 94, 2 deaths). The preschool group (13–60 months) did not reach significance (aOR 4.21, 95% CI 0.92 to 19.17, *p* = 0.063). ETAT emergency category was not independently significant after adjustment (aOR 3.60, 95% CI 0.75 to 17.35, *p* = 0.110). Lower socioeconomic status was associated with mortality in bivariable analysis within the complete case regression subset (cOR 3.98, 95% CI 1.21 to 13.06, *p* = 0.023) but not independently in the multivariable model (aOR 3.03, 95% CI 0.88 to 10.50, *p* = 0.080).

A pre-specified sensitivity analysis was conducted using collapsed referral categories and the full analytic cohort (*n* = 674, 67 events, EPV = 6.7). Referral sources were grouped as: self-presented (reference); lower tier (district/municipal, CHAG, polyclinic, health centre, maternity, and private hospitals combined, *n* = 356); secondary/regional hospitals (*n* = 42); and tertiary institutions (*n* = 27). The same age, ETAT, and SES covariates were retained. No referral tier was independently associated with inpatient mortality. Referral from secondary/regional hospitals was not independently significant (aOR 2.15, 95% CI 0.76 to 6.07, *p* = 0.150), and collapsed lower-tier referral was not associated with mortality (aOR 1.19, 95% CI 0.63 to 2.25, *p* = 0.590). ETAT emergency category was independently associated with inpatient mortality (aOR 3.82, 95% CI 1.04 to 13.97, *p* = 0.043), as was lower socioeconomic status (aOR 4.30, 95% CI 1.49 to 12.44, *p* = 0.007). Age group associations were attenuated to non-significance in the sensitivity model. Both models are exploratory; no estimate should be treated as a confirmed finding.

## DISCUSSION

### Principal findings

This prospective cohort study characterised referral patterns, triage acuity, diagnostic case mix, and inpatient outcomes among children presenting to a national tertiary paediatric emergency unit in Ghana. The descriptive findings represent the primary contribution of this study. The multivariable analysis is exploratory; the primary model had an EPV of 4.2 and all estimates should be treated as such. In the primary analysis, no referral tier was independently associated with inpatient mortality after adjustment. Referral from secondary/regional hospitals showed a borderline non-significant association (aOR 3.09, *p* = 0.058). School going children and adolescents were independently associated with higher odds of death than infants. ETAT category and socioeconomic status were not independently significant in the primary model but were in the sensitivity analysis; this discrepancy is addressed in the subsections below.

### Descriptive findings

A referral rate of 63.0% is at the higher end of the range reported for tertiary paediatric emergency units in SSA. At a comparable facility in Mozambique, 96.4% of presentations were direct attendances without referral from a lower-level facility, and fewer than 10% met ETAT emergency criteria. [17] The higher referral rate and emergency proportion at KBTH are consistent with Ghana’s multi-tier referral system, which channels severely ill children to the national tertiary level. A further 37.0% presented without referral; direct access to tertiary facilities without prior lower-tier assessment is a pattern documented across SSA. [5,8,9,24]

Blood and haematological disorders accounted for 17.0% of presentations, consistent with the high burden of sickle cell disease in Ghana, where approximately 2% of newborns are diagnosed annually and crisis presentations include vaso-occlusive episodes, and Anaemia. [18] Respiratory (17.2%) and digestive presentations (16.4%) are among the most frequently reported diagnostic domains at tertiary paediatric inpatient facilities in SSA, where pneumonia and acute diarrhoeal illness remain the leading infectious causes of under-five morbidity. [19] Both communicable and non-communicable diseases were represented in the cohort, consistent with the epidemiological transition in hospitalised children documented in Ghana. [4]

Inpatient mortality of 33.3% in neoplastic presentations and 31.0% in circulatory presentations most likely reflects the absence of treatment capacity rather than failure of clinical care at the individual level. In SSA, most children with cancer are diagnosed at a stage when curative treatment is no longer possible. [20] In Ghana, fewer than 1% of children with congenital heart disease receive diagnosis and treatment within the first two years of life. [21] Across SSA, fewer than 5% of children requiring cardiac surgery receive it. [22]

### Referral source and mortality

Referral from secondary/regional hospitals showed a borderline non-significant association with inpatient mortality after adjustment (aOR 3.09, 95% CI 0.96 to 9.90, *p* = 0.058). Several caveats apply. The secondary/regional hospital subgroup was small (*n* = 39, 6 mortality events in the regression subset), and the point estimate is statistically unstable. The finding did not reach the pre-specified significance threshold and should not be cited as a precise effect estimate.

Three explanations are plausible. Secondary/regional hospitals may refer the most severely ill children who cannot be managed at that tier; in Ghana, referral decisions at lower-level facilities are typically driven by illness severity, disease complications, and non-response to initial treatment. [12,24] If so, the association reflects disease severity at the point of referral rather than a care failure. Alternatively, delayed recognition of deteriorating illness or late referral at the secondary/regional tier may result in children arriving at KBTH in a worse clinical state than if transferred earlier. The ETAT categorisation on admission reflects the child’s acuity at arrival but cannot determine whether deterioration occurred at the referring facility or during transit. Late presentation at tertiary paediatric emergency units in SSA has been associated with increased disease severity and mortality on admission. [5,8] A third explanation is unmeasured differences in disease severity not captured by the available covariates. The study cannot distinguish between these explanations.

Referral from district and municipal hospitals and CHAG facilities was not independently significant (aOR 2.03, 95% CI 0.89 to 4.61, *p* = 0.091). A direction-consistent pattern across facility tiers is present, but none reached significance.

The lower inpatient mortality among self-presenting children (7.6%) and private hospital referrals (5.9%) compared with public-sector hospital referrals should be interpreted with caution. Self-presenting children had a lower ETAT emergency category rate on arrival (27.6% vs 32.7% among referred children), suggesting a higher proportion of less severely ill cases. Private hospital referrals may reflect earlier presentation, better pre-referral stabilisation, or lower case complexity rather than a difference in care quality.

### ETAT triage acuity and mortality

ETAT emergency category showed a consistent unadjusted association with inpatient mortality in both the full cohort (cOR 4.42, 95% CI 1.31 to 14.85) and the complete case regression subset (*n* = 613; cOR 4.80, 95% CI 1.10 to 20.85, *p* = 0.036). It was not independently significant in the primary multivariable model (aOR 3.60, 95% CI 0.75 to 17.35, *p* = 0.110) but was independently associated in the sensitivity analysis (aOR 3.82, 95% CI 1.04 to 13.97, *p* = 0.043). The descriptive data show a consistent ETAT-mortality gradient: 21.6% mortality in the emergency category compared with 4.6% in the priority category. These data support the prognostic relevance of ETAT classification in this population. Non-significance in the primary model may reflect insufficient statistical power at EPV = 4.2, though a genuine absence of effect cannot be excluded.

### Socioeconomic status and inpatient mortality

The bivariable association between lower SES and inpatient mortality (cOR 3.98, 95% CI 1.21 to 13.06, *p* = 0.023) is consistent with the relationship between household wealth and child mortality documented across SSA, where lower wealth is a primary contributor to variability in under-five mortality rates. Lower SES did not retain independent significance in the primary multivariable model (aOR 3.03, 95% CI 0.88 to 10.50, *p* = 0.080). Data from the CHAIN prospective cohort study of 3,101 children admitted to nine hospitals in SSA and South Asia offer a plausible interpretive framework: structural equation models in that study suggested that socioeconomic factors act on inpatient child mortality indirectly, through clinical and nutritional intermediaries, rather than through a direct independent pathway. [23] In this cohort, referral facility tier is the most plausible intermediary. Lower SES is associated with delayed care-seeking and use of lower-tier facilities in LMIC settings, [9] and a direction-consistent gradient across referral tiers was observed despite no tier reaching significance. When SES and referral tier are both in the model, the shared pathway is removed and the SES coefficient is attenuated. The sensitivity analysis estimate (aOR 4.30, 95% CI 1.49–12.44, *p* = 0.007) is consistent with a genuine SES-mortality relationship. With 51 deaths in 613 complete-case participants, the model EPV was 4.2; limited statistical power may contribute to both the primary model results.

### Age and inpatient mortality

Older age was independently associated with inpatient mortality in the primary model, a direction consistent with the shift in diagnostic case mix across the paediatric age spectrum in SSA: infectious conditions dominate among young children, [1] while neoplastic and non-communicable conditions become more prominent with age and carry higher institutional mortality in settings with limited specialist capacity. [4] In this cohort, 56.7% of neoplastic presentations occurred in children aged 60–119 months, and neoplastic disease had the highest inpatient CFR at 33.3%. Childhood cancer in SSA is predominantly diagnosed in children over five years of age, typically at a stage when curative treatment is no longer possible. [20] This concentration of a high-mortality diagnosis in the school going age group is the most plausible explanation for the aOR of 5.56 for that group. Circulatory presentations, which also carry a high CFR, were distributed across all age groups. Confidence intervals were wide for all age estimates, reflecting the small infant reference group (*n* = 94, 2 deaths); these figures should not be cited as precise effect sizes.

The age-mortality associations should not be interpreted as a direct biological or health system effect of age on the probability of death. In this model, age partly acts as a proxy for the type of condition admitted. Separating the age effect from the diagnostic case mix effect would require including ICD-10 diagnostic domain as a covariate; this was not feasible within the EPV constraints of this cohort and would be a priority in any replication study with larger event numbers.

### Strengths and limitations

The prospective design, consecutive recruitment, and systematic collection of clinical, referral, and demographic variables are the principal strengths of this study. The integration of ETAT data, ICD-10 classification, and referral source within a single cohort enables a characterisation of the emergency care pathway at this facility from referral source through triage acuity and diagnostic case mix to inpatient outcome.

Several limitations apply, particularly to the multivariable analysis. The borderline non-significant association between secondary/regional hospital referral and inpatient mortality rests on *n* = 39 participants and 6 deaths in the regression subset (n = 613, EPV = 4.2), which is insufficient for a stable estimate. All multivariable estimates should be interpreted as exploratory. Penalised regression methods were not used; this limits the reliability of the estimates.

The primary regression excluded 62 participants with incomplete covariate data, and the pattern of exclusion was non-random. Of the 62 excluded participants, 46 (74%) were in the infant age group (≤ 12 months). This concentration is consistent with the known difficulty of obtaining reliable blood pressure measurements in critically ill infants during emergency triage in resource-limited settings. The retained infant subgroup (*n* = 94, 2 deaths, CFR = 2.1%) is therefore a selected, lower-acuity subset; the full-cohort infant CFR was 10.0% (14/140). This inflates the relative aOR estimates for older age groups and should be considered when interpreting the age-group findings.

Children who deteriorated or died during transfer before reaching the PEU are not captured in this cohort. This is a form of survivor bias that may have underestimated the true mortality burden associated with referral from more distant facilities and may have attenuated the observed referral-mortality association.

Presentation timing was available only for referred children and was not included in the multivariable model; its role as a potential mediator of the referral-mortality association remains unexamined.

The single-centre design limits generalisability to other tertiary paediatric emergency settings in Ghana and the wider region. Causal inferences cannot be drawn from these observational data.

### Implications for referral system research

The multivariable findings should be interpreted as hypothesis-generating. If the association between secondary/regional hospital referral and inpatient mortality is confirmed in larger studies, it would justify systematic audit of referral processes and pre-referral stabilisation practices at this tier of the Ghanaian health system. Such confirmation would also support prospective investigation of transfer timing, clinical condition at departure from referring facilities, and whether ETAT-based stabilisation protocols before transfer reduce mortality at the receiving institution. Confirmation requires a multicentre study with adequate statistical power and data on time from referral decision to departure, clinical parameters at the point of referral, stabilisation performed before transfer, and clinical status at arrival.

## CONCLUSIONS

This prospective cohort study characterised referral patterns, ETAT triage acuity, diagnostic case mix, and inpatient outcomes by referral facility tier among 675 children presenting to a national tertiary paediatric emergency unit in Ghana. Of the cohort, 63.0% were referred; 30.8% met ETAT emergency criteria on arrival, with a higher emergency category rate among referred than self-presenting children. Unadjusted inpatient mortality was highest among secondary/regional hospital referrals (16.7%) and district and municipal hospital referrals (16.8%), and highest by diagnosis in neoplastic (33.3%) and circulatory (31.0%) presentations. In the primary multivariable analysis (*n* = 613), no referral tier was independently associated with inpatient mortality. Referral from secondary/regional hospitals showed a borderline non-significant association (aOR 3.09, 95% CI 0.96 to 9.90, *p* = 0.058). School going children and adolescents were independently associated with higher mortality than infants. In the pre-specified sensitivity analysis, ETAT category and lower SES were independently associated with inpatient mortality. All multivariable estimates are exploratory. The descriptive epidemiology of this cohort is the primary contribution of this study.

## Data Availability

Data Availability Statement Manuscript: Referral pathways, ETAT triage acuity, and inpatient outcomes among children presenting to a national tertiary paediatric emergency unit in Ghana: a prospective cohort study Statement for submission system The minimal anonymised dataset supporting the conclusions of this article is available as Supporting Information (S1 Dataset), submitted alongside this manuscript. The dataset contains participant-level data for all 675 enrolled children, with all direct identifiers removed. No external repository deposition has been made at this stage the dataset is provided within the submission package for peer review. If the manuscript is accepted, the corresponding author commits to depositing the dataset in a publicly accessible repository (such as Zenodo or Dryad) prior to publication and providing the persistent DOI in the final version. Justification for non-deposition at submission Public deposition of the full dataset prior to peer review has not been completed for the following reason. The data were collected at a national tertiary hospital (Korle Bu Teaching Hospital, Accra, Ghana) and ethical approval (KBTH-IRB 00073/2017) was granted for use within the scope of this research. The dataset contains clinical, socioeconomic, and diagnostic information from children who are patients at the institution. Although direct identifiers have been removed, the combination of variables (age category, referring facility category, ICD-10 diagnostic code, ETAT category, outcome, Oyedeji SES class) may carry residual re-identification risk in a small population with rare diagnoses. Pre-deposition in a fully open repository without peer review therefore warrants IRB review of the proposed repository terms. This review will be completed prior to publication. Variables included in S1 Dataset The anonymised dataset includes the following variables for each of the 675 participants: Participant identifier (de-identified sequential number) Age group (infant 1-12 months preschool 13-60 months school going 60-119 months adolescent 120-155 months) Sex Socioeconomic status (Oyedeji class: upper I-II middle III lower IV-V) Referral status (referred / self-presented) Referring facility category (self-presented secondary/regional district/municipal/CHAG polyclinic/health centre/maternity home private tertiary) Time from symptom onset to PEU registration in hours (referred children only) ETAT triage category at arrival (emergency priority queue) ICD-10 diagnostic domain (first-level chapter) Inpatient outcome (survived died) Discharge destination (PEU discharge paediatric medical ward surgical ward ICU/HDU special ward absconded) Length of stay in days Blood pressure recorded (yes/no determines inclusion in companion PEWS cohort of n=613) Complete case indicator for primary regression (all covariates present used for n=613 primary model in the revised analysis this corresponds to the same participants as the blood pressure recorded variable above) The following variables were collected but are outside the scope of analyses reported in this manuscript and are not included in S1 Dataset: religion and geographic region of origin. Note on companion manuscripts Three companion manuscripts from the same cohort use a sub-dataset of n=613 participants (those with complete blood pressure data for PEWS score calculation). The n=675 dataset submitted here is the parent dataset. The 62 additional participants (those without blood pressure data, mortality 25.8%) are present only in this dataset and not in the companion PEWS datasets. The S1 Datasets for each companion paper are distinct files derived from the same source data.

## DECLARATIONS

### Ethics approval and consent to participate

Ethical approval: KBTH Institutional Review Board, KBTH-IRB 00073/2017 (with extension). Written informed consent from caregivers; child assent for children aged seven years and above.

### Availability of data and materials

The minimal dataset supporting the conclusions of this article is available as Supporting Information (S1 Dataset). The dataset contains anonymised participant-level data for all 675 enrolled children, with all direct identifiers removed. Age is provided as a categorical variable in the submitted dataset (infant ≤12 months; preschool 1–5 years; school going 5–9 years; adolescent 10–12 years). Age groupings in the multivariable regression (Table 3) were derived from continuous age in months in the original analytical file and differ from this variable. Crude odds ratios in Table 1 were computed in the original SPSS analytical file; marginal differences from the submitted dataset may occur due to variable coding differences. If accepted, the dataset will be deposited in a publicly accessible repository (Zenodo or Dryad) prior to publication and a persistent identifier will be provided in the final version.

### Authors’ contributions

WKAO: conceptualisation, design, data collection, analysis, writing (original draft, review and editing, final approval). LR: supervision, critical revision, and final approval. YA: statistical analysis, review, editing and final approval. TN: statistical analysis, review, editing and final approval. BQG: supervision, critical revision, and final approval. All authors take responsibility for the accuracy of the data and analyses reported.

## Acknowledgements

We thank the children and caregivers who participated in this study. We acknowledge the clinical and nursing staff of the Paediatric Emergency Unit, Department of Child Health, Korle Bu Teaching Hospital, for their support during data collection. We are grateful to the research assistants for their role in prospective data collection and management.

## Companion manuscript statement

Three companion manuscripts from the same prospective cohort have been submitted for publication and are currently under review. A PEWS validation study for mortality prediction is under review at BMJ Paediatrics Open. A PICU eligibility burden analysis is under review at BMC Pediatrics. A prospective observational study comparing the concordance and clinical utility of the Brighton and Bedside PEWS is under review at the African Journal of Emergency Medicine. No Paediatric Early Warning Score data appear in this manuscript. The four papers address non-overlapping research questions with no duplicate content.

## REFERENCES

1. UN Inter-agency Group for Child Mortality Estimation. Levels and trends in child mortality: report 2022 [Internet]. New York: UNICEF; 2022 [cited 2025 Jan]. Available from: https://childmortality.org/

2. Kallah-Dagadu G, Donkor F, Duah M, Yeboah H, Arku D, Lotsi A. Investigation of factors influencing infant mortality at Greater Accra Regional Hospital, Ghana. Biomed Res Int. 2024;2024:6610617. doi:10.1155/2024/6610617.

3. Poulin D, Nimo G, Royal D, Joseph PV, Nimo T, Nimo T, et al. Infant mortality in Ghana: investing in health care infrastructure and systems. Health Aff Sch. 2024;2(2):qxae005. doi:10.1093/haschl/qxae005.

4. Yawson AE, Abuosi AA, Badasu DM, Atobra D, Adzei FA, Anarfi JK. Non-communicable diseases among children in Ghana: health and social concerns of parent/caregivers. Afr Health Sci. 2016;16(2):378–88. doi:10.4314/ahs.v16i2.6.

5. Molyneux EM, Goka BQ. Paediatric emergencies in sub-Saharan Africa. Afr J Emerg Med. 2017;7(Suppl 1):S1–2. doi:10.1016/j.afjem.2017.11.005.

6. Dekker-Boersema J, Hector J, Jefferys LF, Binamo C, Camilo D, Muganga G, et al. Triage conducted by lay-staff and emergency training reduces paediatric mortality in the emergency department of a rural hospital in Northern Mozambique. Afr J Emerg Med. 2019;9(4):172–176. doi:10.1016/j.afjem.2019.05.005.

7. English M, Esamai F, Wasunna A, Were F, Ogutu B, Wamae A, et al. Assessment of inpatient paediatric care in first referral level hospitals in 13 districts in Kenya. Lancet. 2004;363(9425):1948–53. doi:10.1016/S0140-6736(04)16408-8.

8. Simoes EAF, Peterson S, Gamatie Y, Kisanga FS, Mukasa G, Nsungwa-Sabiiti J, et al. Management of severely ill children at first-level health facilities in sub-Saharan Africa when referral is difficult. Bull World Health Organ. 2003;81(7):522–531. doi:10.2471/blt.2003.81.7.522.

9. Clarke-Deelder E, Osei Afriyie D, Nseluke M, Masiye F, Fink G. Health care seeking in modern urban LMIC settings: evidence from Lusaka, Zambia. BMC Public Health. 2022;22(1):1205. doi:10.1186/s12889-022-13549-3.

10. Asamani JA, Chebere MM, Barton PM, D’Almeida SA, Odame EA, Oppong R. Forecast of healthcare facilities and health workforce requirements for the public sector in Ghana, 2016-2026. Int J Health Policy Manag. 2018;7(11):1040–52. doi:10.15171/ijhpm.2018.64.

11. Gyedu A, Baah EG, Boakye G, Ohene-Yeboah M, Otupiri E, Stewart BT. Quality of referrals for elective surgery at a tertiary care hospital in a developing country: an opportunity for improving timely access to and cost-effectiveness of surgical care. Int J Surg. 2015;15:74–78. doi:10.1016/j.ijsu.2015.01.033.

12. World Health Organization. Emergency triage assessment and treatment (ETAT): manual for participants [Internet]. Geneva: WHO; 2005 [cited 2025 Jan]. Available from: https://iris.who.int/handle/10665/43386

13. King C, Dube A, Zadutsa B, Banda L, Langton J, Desmond N, et al. Paediatric Emergency Triage, Assessment and Treatment (ETAT): preparedness for implementation at primary care facilities in Malawi. Glob Health Action. 2021;14(1):1989807. doi:10.1080/16549716.2021.1989807.

14. Oyedeji GA. Socio-economic and cultural background of hospitalised children in Ile-Ife. Niger J Paediatr. 1985;12:111–7.

15. Ghana Statistical Service, Health Research Unit, Ministry of Health, and ORC Macro. Ghana Service Provision Assessment Survey 2002 [Internet]. Calverton, Maryland: Ghana Statistical Service and ORC Macro; 2003 [cited 2025 Jan]. Available from: https://dhsprogram.com/publications/publication-spa6-spa-final-reports.cfm

16. Christian Health Association of Ghana (CHAG). Regional overview of CHAG facilities [Internet]. Accra: CHAG; 2022 [cited 2025 Jan]. Available from: https://chag.org.gh/where-we-serve/regional-overview-of-chag-facilities/

17. Brugnolaro V, Fovino LN, Calgaro S, Putoto G, Muhelo AR, Gregori D, et al. Pediatric emergency care in a low-income country: Characteristics and outcomes of presentations to a tertiary-care emergency department in Mozambique. PLOS ONE. 2020;15(11):e0241209. doi:10.1371/journal.pone.0241209.

18. Asare EK, Olayemi E, Boafor TK, Dei-Adomakoh Y, Mensah E, Afriyie-Mensah JS, et al. Burden of sickle cell disease in Ghana: The Korle-Bu experience. Adv Haematol. 2018;2018:6161270. doi:10.1155/2018/6161270.

19. Owusu DN, Duah HO, Dwomoh D, Alhassan Y. Prevalence and determinants of diarrhoea and acute respiratory infections among children aged under five years in West Africa: evidence from demographic and health surveys. Int Health. 2024;16(1):97–106. doi:10.1093/inthealth/ihad046.

20. Lubega J, Chirande L, Atwine B, Davidson A, Kashaigili HJ, Kanyamuhunga A, et al. Addressing the childhood cancer crisis in sub-Saharan Africa. Lancet Oncol. 2023;24(7):729–732. doi:10.1016/S1470-2045(23)00173-0.

21. Manuel V, Miana LA, Edwin F. Narrative review in pediatric and congenital heart surgery in sub-Saharan Africa: challenges and opportunities in a new era. AME Surg J. 2021;1:26. doi:10.21037/asj-21-34.

22. Leith J, Harik L, An KR, Brashear T, Peck RN, Bhamidipati CM. Estimating congenital cardiac surgical need in Africa using geographic distribution of surgeons. Ann Glob Health. 2025;91(1):28. doi:10.5334/aogh.4692.

23. CHAIN Network. Childhood mortality during and after acute illness in Africa and south Asia: a prospective cohort study. Lancet Glob Health. 2022;10(5):e673–684. doi:10.1016/S2214-109X(22)00118-8.

24. Afrifa-Yamoah E, Nunfam VF, Kwanin BA, Frimpong K. Ecology of emergency care in lower-tier healthcare providers in Ghana: an empirical data-driven Bayesian network analytical approach. Intern Emerg Med. 2024;19(4):1081–96. doi:10.1007/s11739-024-03607-6.

